# Placental PFAS concentrations are associated with perturbations of placental DNA methylation at loci with important roles on cardiometabolic health

**DOI:** 10.1101/2024.05.06.24306905

**Authors:** Todd M. Everson, Neha Sehgal, Dana Boyd Barr, Parinya Panuwet, Volha Yakimavets, Cynthia Perez, Kartik Shankar, Stephanie M. Eick, Kevin J. Pearson, Aline Andres

## Abstract

The placenta is crucial for fetal development, is affected by PFAS toxicity, and evidence is accumulating that gestational PFAS perturb the epigenetic activity of the placenta. Gestational PFAS exposure is can adversely affect offspring, yet individual and cumulative impacts of PFAS on the placental epigenome remain underexplored. Here, we conducted an epigenome-wide association study (EWAS) to examine the relationships between placental PFAS levels and DNA methylation in a cohort of mother-infant dyads in Arkansas. We measured 17 PFAS in human placental tissues and quantified placental DNA methylation levels via the Illumina EPIC Microarray. We tested for differential DNA methylation with individual PFAS, and with mixtures of multiple PFAS. Our results demonstrated that numerous epigenetic loci were perturbed by PFAS, with PFHxS exhibiting the most abundant effects. Mixture analyses suggested cumulative effects of PFOA and PFOS, while PFHxS may act more independently. We additionally explored whether sex-specific effects may be present and concluded that future large studies should explicitly test for sex-specific effects. The genes that are annotated to our PFAS-associated epigenetic loci are primarily involved in growth processes and cardiometabolic health, while some genes are involved in neurodevelopment. These findings shed light on how prenatal PFAS exposures affect birth outcomes and children’s health, emphasizing the importance of understanding PFAS mechanisms in the in-utero environment.

## Introduction

Per- and polyfluoroalkyl substances (PFAS) are man-made chemicals that are ubiquitously used in consumer products, food packaging, and firefighting foams. These chemicals are of significant environmental concerns due to their pervasiveness in everyday products and their persistence in the environment and human biomatrices [1]. Human exposure to PFAS is linked to several health outcomes, including multiple cardiometabolic conditions, and adverse effects during pregnancy (e.g., birth and developmental outcomes) [2]. Several mechanisms could explain these intergenerational effects, including direct fetal exposure from maternal circulation or by disrupting maternal physiology including placental development and function. Some quantity of PFAS from maternal circulation pass through the placenta to reach many fetal organ systems, and PFAS bioaccumulate in the placenta throughout gestation [3]. PFAS accumulation in the placenta may have consequences on placental function and moderation of the *in-utero* environment.

A recent review Szilagyi et al (2020) highlighted a likely role for PFAS interactions with peroxisome proliferator-activated receptors (PPARs), which may impact placental functions [4]. PPARs are nuclear receptors that broadly regulate lipid, hormone and glucose metabolism [5], and are associated with placentation and trophoblast differentiation [4]. PPAR activities in the placenta serve many roles, and influences the expression of multiple genes and pathways involved in placental development [6].

Additionally, epigenetic alterations, such as changes in DNA methylation, may impact placental function, affect birth outcomes and children’s health, since other persistent organic pollutants (POPs) have such effects [7]. A few recent studies have begun to explore the impact of prenatal PFAS on the human placental epigenome. Wang et al (2023) used a targeted gene approach and global methylation analysis, to show that placental PFAS levels were negatively associated LINE-1 methylation and with CpGs in the nuclear receptor subfamily 3 group C member 1 (*NR3C1*) gene; and that PFAS associations with placental methylation and birth outcomes may be modified by sex [8]. An epigenome-wide association study (EWAS) of several persistent organic pollutants in maternal plasma, including PFAS, found that several individual PFAS were associated with differences in DNA methylation levels of 39 CpG sites [9].

Another study examined relationships between maternal plasma PFAS concentrations with placental DNAm differences using reduced representation bisulfite sequencing in a sub-sample (n=16), then demonstrated more statistically robust associations with three of these five candidates in the full cohort (n=345) [10]. Thus, to date, studies of the impacts of gestational PFAS exposure on the human placental methylome have primarily utilized maternal plasma as an exposure biomarker, or when placental PFAS levels were quantified, they were only examined in association with targeted genes and features.

Given that the placenta is likely affected by PFAS toxicity, and individual PFAS have been shown to be associated with DNA methylation differences in a handful of placental epigenomic studies, there is a need to further characterize the cumulative impacts of PFAS in the placenta on the placental epigenome. Such studies which better reflect real world PFAS exposure patterns can help to elucidate the mechanisms by which PFAS exert their effects on placental function and the *in-utero* environment. Here, we performed an EWAS of the relationships between placental PFAS concentrations with differences in DNA methylation. We first examined these relationships for each individual PFAS compound that was uniformly detected in our sample, then examined the associations with the overall mixture of PFAS on DNA methylation, and last explored whether there was evidence of sex-specific effects.

## Methods

### Study Population

This study was performed as part of the Glowing Study in Little Rock, Arkansas, which recruited pregnant women prior to gestational week 10, and enrollment occurred between 2010 and 2014 (www.clinicaltrials.gov, ID #NCT03281850) [11, 12]. This longitudinal observational cohort consisted of 300 second parity dyads, from singleton pregnancies, with mothers who were at least 21 years of age.

Exclusion criteria included self-reported pre-existing medical conditions, sexually transmitted infections, medical complications, reported smoking or alcohol use during pregnancy, or medication use during pregnancy known to influence fetal growth (e.g. gestational diabetes, thyroid disease, pre-eclampsia) and conceptions aided with fertility treatment. Infants included in the analyses had no medical conditions and required no medications known to influence growth or development of the fetus. This study included all dyads where placental samples were collected, and where PFAS and DNA methylation quantification passed quality control (n=151). All study participants provided informed, written consent and the study was approved by the University of Arkansas for Medical Sciences Institutional Review Board.

### Placental Collection and Processing

Placentas were collected within 30 minutes of delivery and processed within 2 hours, those with severe placental pathologies were excluded. Placental tissues from the villous core, maternal and fetal sides were separately collected from 6 random sites (∼1 sq. in) and washed thrice in ice-cold PBS to remove maternal blood. Samples were then flash-frozen. For comprehensive representation of the entire placenta, ∼1 g of pooled tissue from all six collection sites (each compartment separately) was frozen in liquid nitrogen and pulverized. Tissue samples were pooled from all regions of the villous core and the same pooled pulverized sample was used for all -omics and PFAS analysis.

### Placental PFAS Quantification

Placental samples were randomized prior to analysis to reduce analytical batch effects. Powdered placental tissue (0.2-0.7mg per sample), was spiked with isotopically labeled analogues of target PFAS, then homogenized in methanol. The 17 target PFAS include: perfluorohexanoic acid (PFHxA), perfluorohexane sulfonic acid (PFHxS), perfluoroheptanoic acid (PFHpA), perfluorooctanoic acid (PFOA), perfluorooctane sulfonic acid (PFOS), perfluorooctane sulfonamide (PFOSA), n- methylperfluoro-1-octanesulfonamidoacetic acid (MePFOSAA), perfluorononanoic acid (PFNA), perfluorodecanoic acid (PFDA), perfluorodecane sulfonic acid (PFDS), perfluoroundecanoic acid (PFUnDA), perfluorododecanoic acid (PFDoDA), perfluoropentanoic acid (PFPeA), n-ethylperfluoro-1- octanesulfonamidoacetic acid (EtPFOSAA), hexafluoropropyleneoxide dimer acid (HFPO-DA; Gen X), perfluoroheptane sulfonic acid (PFHpS), and perfluorobutanesulfonic acid (PFBS). The homogenized samples were centrifuged, and the methanol layer was separated from the placenta tissue. The methanolic fraction was then evaporated to almost dryness and mixed with a mixture of methanol: 0.1M formic acid (40:60, v/v). The samples were introduced onto a Strata RP on-line extraction column (2.1 x 20 mm). The on-line extraction column was washed with 0.1 M formic acid: acetonitrile (90:10, V/V). Then, the target analytes were transferred to a Betasil C18 analytical column (6 x 150 mm) for chromatographic separation. The target PFAS were analyzed using high-performance liquid chromatography-tandem mass spectrometry (LC-MS/MS) with electrospray ionization. The analyte concentrations were calculated using isotope dilution calibration. The unknown samples were analyzed alongside a replicate of solvent-based blanks for background evaluation and quality control materials (2 levels) for precision and accuracy assurance.

Two samples that had insufficient quantity of tissue to perform quantification were excluded from further analyses, yielding a sample size of 151 for analyses of placental PFAS levels. Concentrations of five PFAS (PFHxS, PFOS, PFOA, PFNA, and PFDA) were detectable in over 70% of placental samples. All downstream analyses focused on these five PFAS. Concentrations below the limit of detection (LOD) were imputed via LOD/√2 and data were natural log-transformed prior to performing statistical analyses.

### Placental DNA methylation Quantification

DNA extraction was performed using PureLink Genomic DNA isolation reagents (Thermo Fisher). DNA was quantified with Qubit dsDNA quantification assays (Thermo Fisher, Waltham, MA, USA), then aliquoted into standardized concentrations to allow for a total mass of 500 ng of DNA.

Samples were randomly distributed across 96-well plates, rows, and chips to reduce the potential for batch effects. The Emory University Integrated Genomics Core performed bisulfite conversion using the EZ DNA Methylation Kit (Zymo Research, Irvine, CA) and measured DNAm throughout the genome using the Illumina MethylationEPIC Beadarray (Illumina, San Diego, CA) following the manufacturer’s protocol. All 153 placental DNA samples passed quality control, with no samples yielding more than 1% or of probes with detection p values > 0.05. Functional normalization [13] and beta-mixture quantile (BMIQ) normalization were performed [14] to normalize technical differences on the arrays and correct for different distributions between type-I and type-II probe designs. Then probes on the X and Y chromosomes, those that had single nucleotide polymorphisms (SNP) within the binding region, and those that could cross-hybridize to other regions of the genome [15] were excluded.

### Statistical Analyses

All statistical analyses were performed in RStudio (version 4.3.0 or above). Frequencies and arithmetic means and standard deviations (SDs) were used to describe the distribution of demographic variables. Geometric means and SDs and selected percentiles were used to summarize PFAS concentrations. Violin plots were used to visualize the distributions of natural log transformed PFAS. Spearman correlation coefficients were used to determine the relation between each placental PFAS.

Prior to conducting the epigenome wide association study (EWAS), we performed two unsupervised data reduction steps to reduce multiple testing burden and increase power. First, we utilized CoMeBack to identify proximal CpG sites that exhibit highly correlated methylation patterns and clustered these CpGs into co-methylated regions (CMRs) [16]. For each CMR, principal component analysis was performed and the first component was used as a summary measure of the of DNAm within each CMR. This step identified 57,146 CMRs representing the DNAm of 158,924 CpG sites. Many CpG sites that were not clustered in CMRs and these were retained as individual CpG sites, for a total of 599,048 epigenetic loci (CpGs + CMRs). Next, since CpGs with low variability are more prone to be affected by measurement error and may yield less reproducible findings [17], we excluded CpGs or CMRs with low standard deviation (SD < 0.02). After performing these data reduction steps, 365,376 epigenetic loci were available for analyses – 49,470 CMRs and 315,906 individual CpGs.

We conducted five EWAS to characterize the relationships between DNAm levels and the concentrations of five individual PFAS chemicals in placental tissue. We used robust linear regression models from the MASS package which implement M-estimation with Huber and bisquare weighting to reduce the influence of outliers[18], to regress DNA methylation beta values on natural log transformed PFAS concentrations. All models were adjusted for technical covariates and potential confounders that were determined *a priori* based on a directed acyclic graph (DAG, **Supplemental Figure S1**). All models were adjusted for maternal BMI (normal weight, overweight/obesity), maternal age at delivery, gestational age at birth, maternal educational attainment, infant sex, DNA methylation batch, and proportions of placental cell types (trophoblasts, stromal cells, Hofbauer cells, endothelial cells, nRBCs, and syncytiotrophoblasts), which were estimated via the *planet* package in R [19]. We evaluated genomic inflation factors and QQ-plots for evidence of inflation and calculated false-discovery rate (FDR) for each PFAS EWAS, considering those associations with FDR q-values < 0.05 as statistically significant.

We then used secondary analyses to evaluate whether there was a mixture effect on DNA methylation from simultaneous exposure to multiple PFAS congeners in the placenta. For this, we performed a targeted analysis of the CpGs that were significantly associated with any one of the PFAS congeners from the primary EWAS. For these analyses we applied quantile g-computation, a parametric generalized linear modelling approach that performs g-computation [20]. This method estimates the overall effect of the placental PFAS mixture by simultaneously increasing all concentrations by one quartile, while controlling for the same potential confounders and technical factors described above. In addition to estimating the overall mixture effect, each PFAS component in the mixture is assigned a partial positive or a partial negative weight, regardless of the direction and magnitude of the overall mixture effect. All weights in the same direction sum to 1, indicating the relative contribution of an individual PFAS towards that direction of effect on methylation for a given loci. Because the weights represent relative contributions for only a particular direction of effect, the weights in a particular direction should only be compared and be evaluated separately for each epigenetic loci. We performed two quantile g-computation analysis to quantify the effect of placental PFAS exposure on each loci. In our first approach, we did not allow for interaction between individual PFAS. In our second approach, we allowed for the model to include all two-way interaction terms using bootstrapping.

Additionally, because emerging evidence suggests that developmental exposure to PFAS may have sexually dimorphic effects on some outcomes [21], we explored whether there was any evidence of sex-specific associations from the individual PFAS or from the PFAS mixture on DNAm. For these analyses, we performed the untargeted EWAS for each of the 5 PFAS among females (N=63) and males (N=88) separately. These models had the same covariate adjustments as above but did not include sex as a covariate.

Last we performed functional enrichment analyses to test whether the sets of epigenetic loci that we identified in our EWAS were components within broader biological processes or functions. We used the *missMethyl* package in R to test for gene ontology (GO) term and Kyoto Encyclopedia of Genes and Genomes (KEGG) pathway enrichment, since this method corrects for probe number bias [22].

## Results

### Study Population

This study sample included mother-infant dyads from the Glowing Study for whom placental DNAm and placental PFAS were quantified. As part of enrollment criteria, all mothers were self-reported non-smokers and were second parity. The study sample (n=151) primarily represented healthy pregnancies and birth outcomes (**Table 1**). The mean maternal age at the time of delivery was 30.5 years (SD = 3.42 years) and most of the mothers had a college degree (66.9%) and self-identified as White (86.1%). The 151 infants included 63 females and 88 males, who were born with an average gestation of 39.3 weeks (range 36.4 – 41.4 weeks).

**Table 1:** Distribution of demographic variables in the Glowing study, 2010-2014.

|  | Overall<br>(N=151) |
| --- | --- |
| **Infant Sex** |  |
| Female | 63 (41.7%) |
| Male | 88 (58.3%) |
| **BMI Enrollment Category** |  |
| Pregnant women with normal weight | 74 (49.0%) |
| Pregnancy women with overweight or obesity | 77 (51.0%) |
| **Maternal Race** |  |
| White | 130 (86.1%) |
| Non-White | 21 (13.9%) |
| **Maternal Educational Attainment** |  |
| College Degree | 101 (66.9%) |
| No College Degree | 50 (33.11%) |
| **Maternal Age at Delivery (years)** |  |
| Mean (SD) | 30.5 (3.42) |
| Median [Min, Max] | 30.2 [22.5, 42.4] |
| **Gestational Age at Birth (weeks)** |  |
| Mean (SD) | 39.3 (0.85) |
| Median [Min, Max] | 39.3 [36.4, 41.4] |

### Placental PFAS Quantification

Concentrations of five PFAS were detectable in over 70% of placental samples: PFHxS, PFOS, PFOA, PFNA, and PFDA. As displayed in **Table 2**, PFOS was detected at the highest rate and had the highest average concentrations, while PFOA and PFHxS had the second and third highest concentrations. Data were natural log-transformed prior to downstream analyses to deal with the right skewed distributions but some distributions for LOD-imputed PFAS remain non-normal (**Supplemental Figure S2**). The placental concentrations of these PFAS were moderately correlated with each other (**Supplemental Figure S3**), with the strongest pairwise correlation between PFNA and PFDA (Spearman rho = 0.71), the weakest correlation between PFHxS and PFDA or PFNA (Spearman rho = 0.30), and PFHxS generally exhibiting lower correlations with other PFAS.

**Table 2:** Distribution of placental per- and polyfluoroalkyl substances levels (ng/g) in the Glowing study, 2010-2014.

|  | Above<br>Limit of<br>Detection<br>(%) | N | Geometric<br>Mean | Geometric<br>Standard<br>Deviation | Percentiles |  |  |  |  |
| --- | --- | --- | --- | --- | --- | --- | --- | --- | --- |
|  |  |  |  |  | 5% | 25% | 50% | 75% | 95% |
| **PFHxS** | 83.4 | 151 | 0.05 | 2.82 | 0.01 | 0.04 | 0.05 | 0.08 | 0.23 |
| **PFOA** | 74.8 | 151 | 0.05 | 2.87 | 0.01 | 0.01 | 0.07 | 0.11 | 0.24 |
| **PFNA** | 98.0 | 151 | 0.05 | 1.86 | 0.02 | 0.03 | 0.05 | 0.08 | 0.12 |
| **PFOS** | 98.7 | 151 | 0.38 | 2.01 | 0.12 | 0.26 | 0.43 | 0.58 | 0.97 |
| **PFDA** | 68.9 | 151 | 0.025 | 2.48 | 0.01 | 0.01 | 0.04 | 0.05 | 0.07 |

### Epigenome-Wide Association Study (EWAS) of Placental PFAS

We performed five EWAS, to estimate differences in DNA methylation across 365,376 epigenetic loci. Genomic inflation factors and QQ plots for each EWAS indicated minimal evidence of inflation (**Supplemental Figure S4**). We identified differential methylation at 23 epigenetic loci (FDR q-values < 0.05; **Figure 1**) with PFHxS having the greatest number of associations (11 loci), followed by PFNA (5 loci), PFOS (4 loci), PFOA (2 loci), and PFDA (1 loci) (**Table 3**). While the loci that were associated each of these five PFAS differed, the parameter estimates for their associations were very highly correlated (**Supplemental Figure S5**). Parameter estimates for PFOS and PFNA exhibited the strongest correlations (Spearman rho = 0.93), while the parameter estimates for PFHxS with all other PFAS were the weakest, but still moderate-to-strong correlations (rho range 0.71 – 0.79). The overall EWAS output will be made publicly available upon publication.

**Figure 1:**
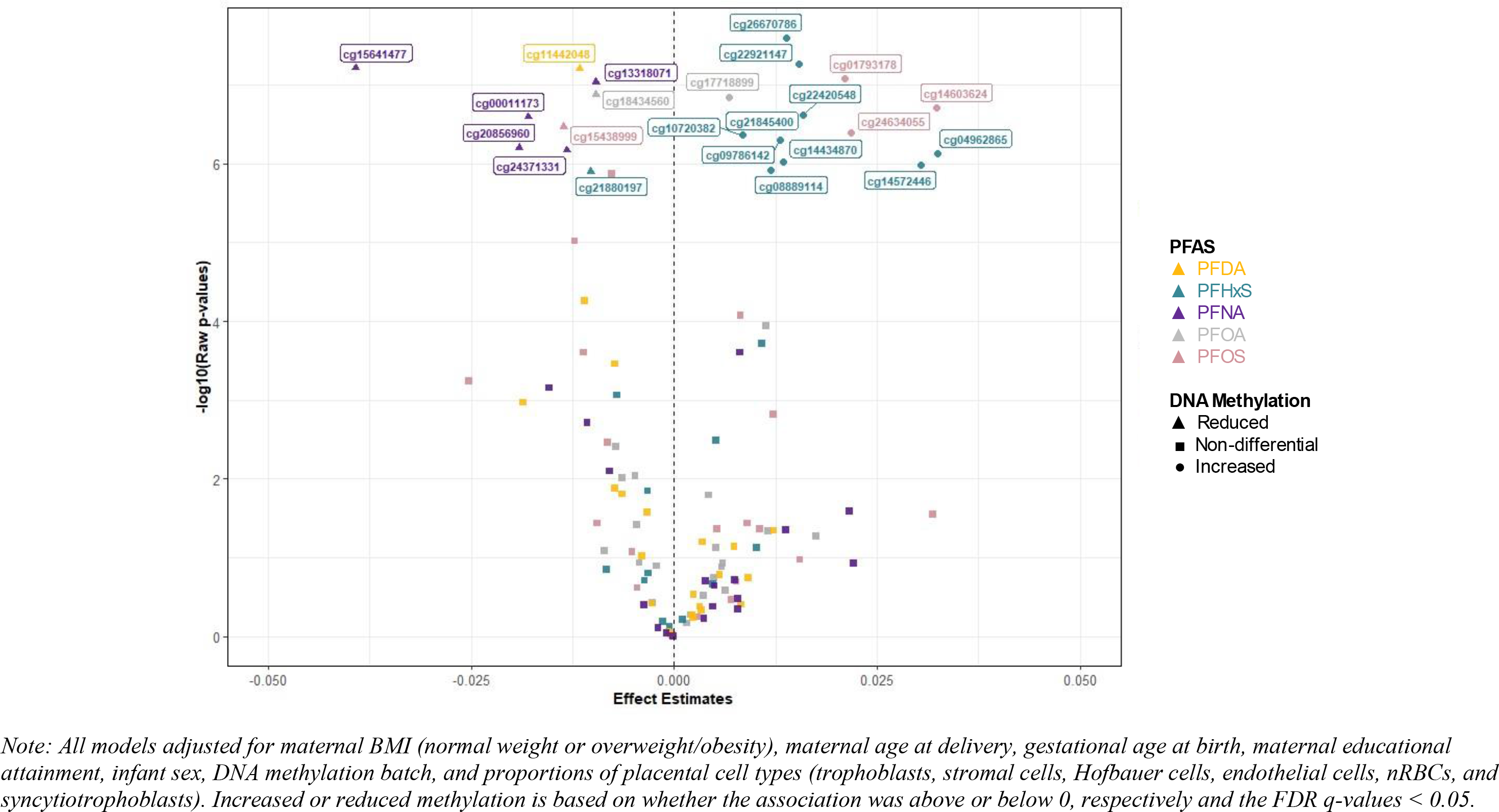
Volcano plot for the associations between increasing concentrations of placental per- and polyfluoroalkyl substances (PFAS) levels with placental DNA methylation levels for the 23 CpGs that were significantly impacted by any one of the five PFAS in the Glowing study (N=151), 2010-2014.

**Table 3:** Placental EWAS results for PFOS, PFOA, PFHxS, PFDA, and PFNA, for the 23 loci that were associated at least one PFAS; Beta represents the estimated differential methylation with a 1-ln increase in PFAS and text is used to highlight those associations that yielded FDR q-values (q-val.) < 0.05; Loci that were CMRs list tiple CpG IDs.

| Epigenetic Loci | PFOS |  | PFOA |  | PFHxS |  | PFDA |  | PFNA |  |
| --- | --- | --- | --- | --- | --- | --- | --- | --- | --- | --- |
|  | Beta | q-val. | Beta | q-val. | Beta | q-val. | Beta | q-val. | Beta | q-val. |
| cg01793178 | 0.0211 | **0.03** | 0.0049 | 1.00 | 0.0108 | 0.22 | -0.0007 | 0.99 | 0.0138 | 1.00 |
| cg15438999 | -0.0136 | **0.04** | -0.0072 | 0.94 | -0.0037 | 0.85 | -0.0111 | 0.31 | -0.0154 | 0.80 |
| cg24634055 | 0.0218 | **0.04** | 0.0016 | 1.00 | 0.0048 | 0.86 | 0.0074 | 0.83 | 0.0048 | 1.00 |
| cg14603624 | 0.0323 | **0.04** | 0.0063 | 1.00 | 0.0102 | 0.76 | 0.0123 | 0.79 | 0.0216 | 1.00 |
| cg18434560 | -0.0122 | 0.18 | -0.0097 | **0.03** | -0.0070 | 0.33 | -0.0039 | 0.85 | -0.0107 | 0.89 |
| cg17718899 | 0.0082 | 0.38 | 0.0067 | **0.03** | 0.0051 | 0.46 | 0.0035 | 0.82 | 0.0081 | 0.61 |
| cg00011173 | -0.0111 | 0.46 | -0.0046 | 1.00 | 0.0011 | 0.96 | -0.0064 | 0.69 | -0.0180 | **0.03** |
| cg24371331 | -0.0082 | 0.77 | -0.0048 | 0.99 | -0.0006 | 0.97 | -0.0073 | 0.38 | -0.0132 | **0.05** |
| cg15641477 | -0.0253 | 0.54 | -0.0086 | 1.00 | -0.0083 | 0.82 | -0.0186 | 0.46 | -0.0393 | **0.02** |
| cg20856960; cg22399868 | -0.0095 | 1.00 | -0.0043 | 1.00 | -0.0014 | 0.96 | -0.0073 | 0.68 | -0.0191 | **0.05** |
| cg13318071; cg18107425 | -0.0077 | 0.10 | -0.0022 | 1.00 | -0.0032 | 0.61 | -0.0033 | 0.75 | -0.0096 | **0.02** |
| cg11442048 | -0.0052 | 1.00 | -0.0064 | 0.99 | -0.0032 | 0.83 | -0.0116 | **0.02** | -0.0080 | 1.00 |
| cg08889114 | 0.0029 | 1.00 | 0.0060 | 1.00 | 0.0119 | **0.04** | 0.0023 | 0.97 | -0.0001 | 1.00 |
| cg21845400 | -0.0002 | 1.00 | 0.0036 | 1.00 | 0.0148 | **0.02** | -0.0004 | 1.00 | -0.0020 | 1.00 |
| cg09786142 | 0.0122 | 0.66 | 0.0113 | 0.55 | 0.0130 | **0.03** | 0.0056 | 0.89 | 0.0074 | 1.00 |
| cg22420548 | 0.0070 | 1.00 | 0.0059 | 1.00 | 0.0159 | **0.02** | 0.0034 | 0.96 | 0.0079 | 1.00 |
| cg14572446 | 0.0155 | 1.00 | 0.0116 | 1.00 | 0.0303 | **0.04** | 0.0091 | 0.90 | 0.0079 | 1.00 |
| cg14434870 | 0.0077 | 1.00 | -0.0005 | 1.00 | 0.0134 | **0.04** | 0.0032 | 0.96 | -0.0010 | 1.00 |
| cg04962865 | 0.0319 | 0.99 | 0.0175 | 1.00 | 0.0325 | **0.04** | 0.0083 | 0.95 | 0.0221 | 1.00 |
| cg26670786 | 0.0090 | 1.00 | 0.0052 | 1.00 | 0.0138 | **0.01** | 0.0022 | 0.97 | 0.0050 | 1.00 |
| cg10720382 | 0.0053 | 1.00 | 0.0042 | 1.00 | 0.0084 | **0.03** | 0.0024 | 0.93 | 0.0039 | 1.00 |
| cg22921147 | 0.0105 | 1.00 | 0.0020 | 1.00 | 0.0153 | **0.01** | -0.0002 | 1.00 | 0.0037 | 1.00 |
| cg21880197 | -0.0045 | 1.00 | -0.0026 | 1.00 | -0.0103 | **0.04** | -0.0027 | 0.95 | -0.0037 | 1.00 |

**Table 4:**
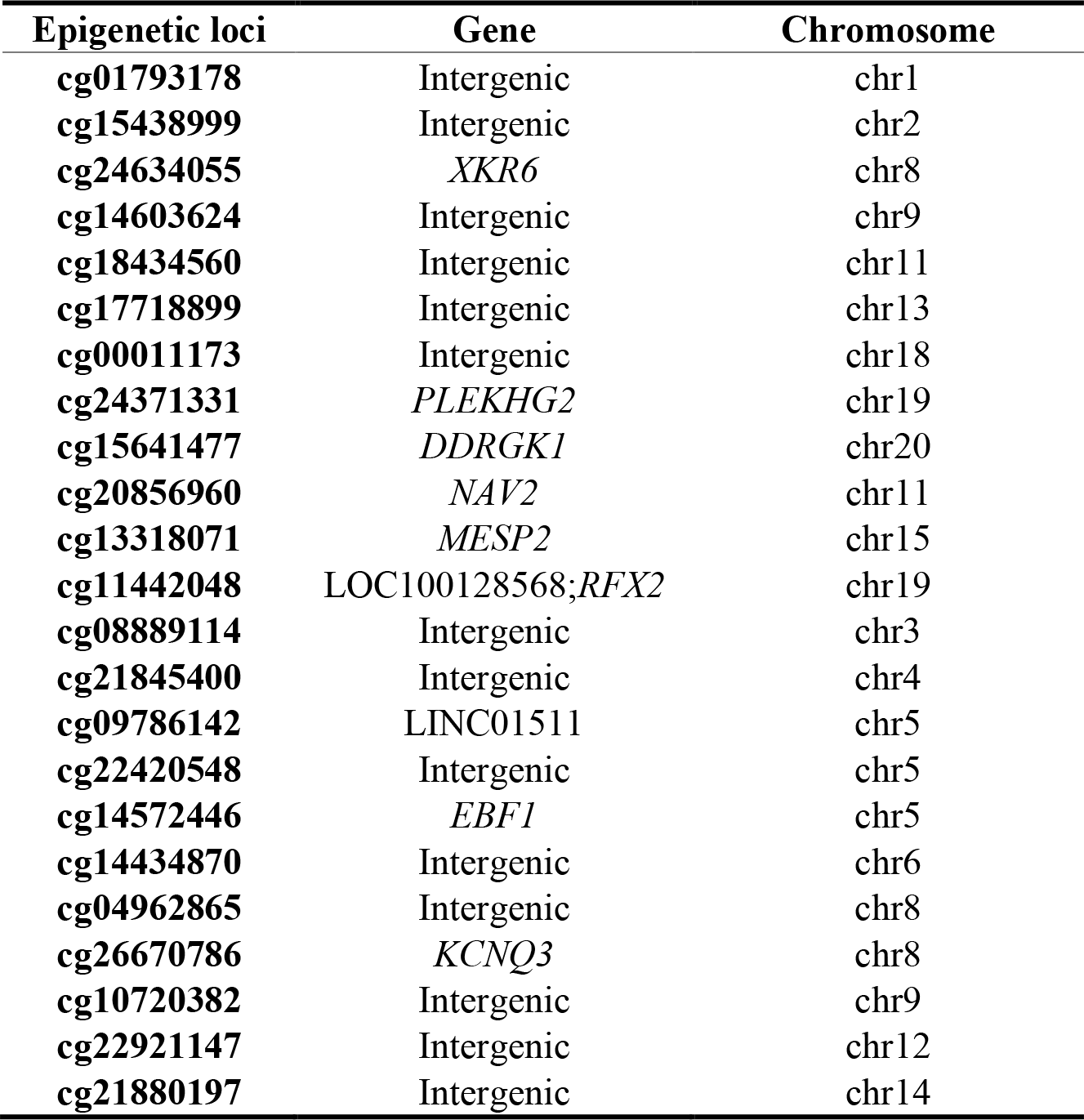
Genomic annotations for 23 CpGs associated with any of PFOS, PFOA, PFHxS, PFDA, or PFNA.

### Secondary PFAS Mixtures Analyses

Since our findings above show that PFAS concentrations in the placenta were moderately correlated with each other, and the 23 loci that were associated with any PFAS exhibited correlated effects on DNAm, we hypothesized that PFAS may not have independent effects on DNAm but instead may have synergistic or cumulative effects. Thus, we examined whether placental DNAm levels at these 23 loci were significantly impacted by simultaneous exposure to PFAS. Using quantile g-computation, we found that overall, a simultaneous increase in all five PFAS by one quartile was associated with reduced methylation levels at 6 loci and with increased methylation levels at 6 loci in the placental tissue (**Figure 2** and **Table S1**). Partial positive and negative weights from quantile g-computation show the proportion of the effect in a particular direction associated with each PFAS. Some of the mixture results were similar to the EWAS results for each PFAS, where PFHxS greatly contributed to the positive effects while PFOS, PFOA, and PFNA primarily contributes to the negative direction (**Figure 3**).

**Figure 2:**
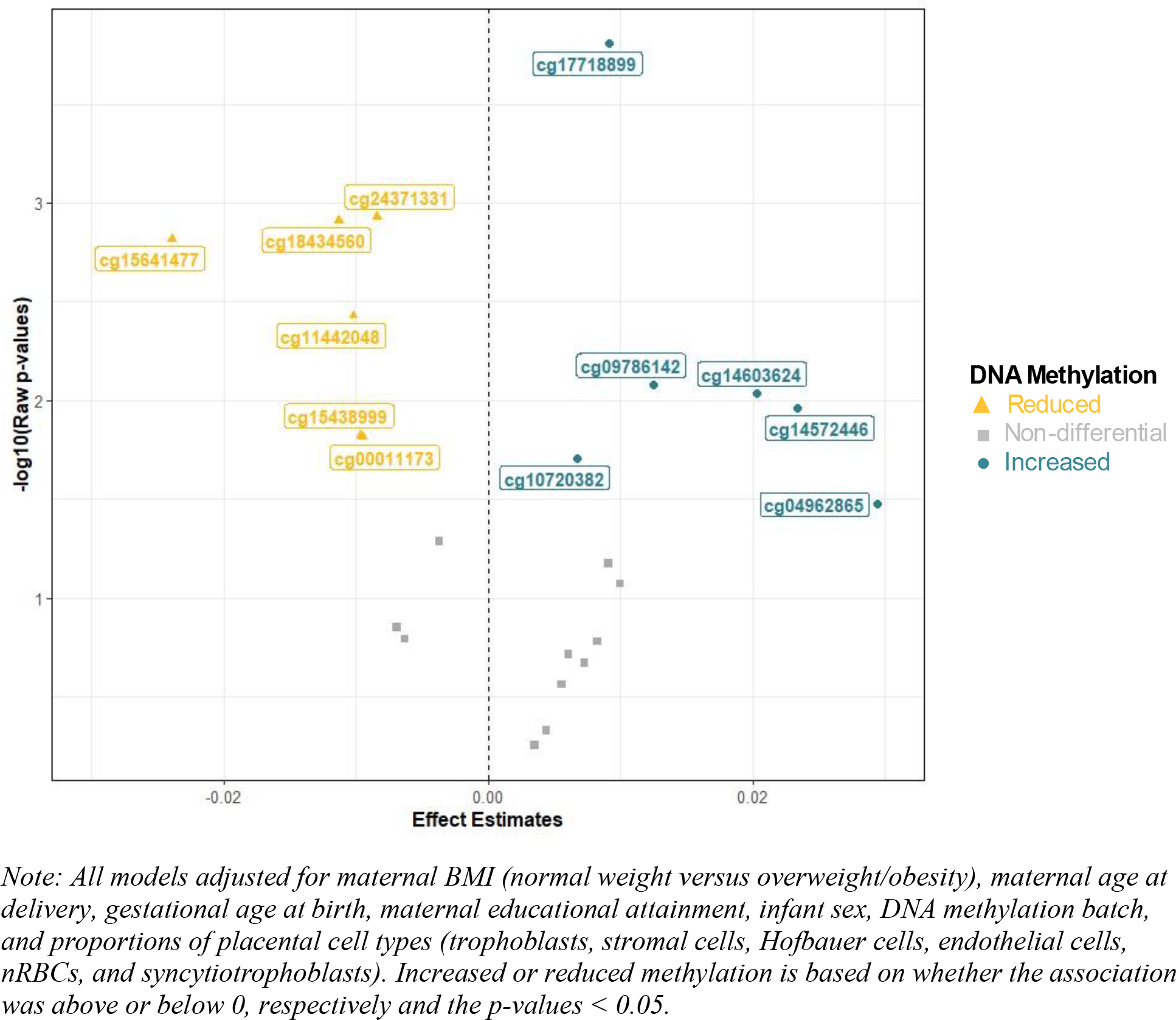
Volcano plots for the mixture effect of placental per- and polyfluoroalkyl substances (PFAS in ng/g) exposure on placental DNA methylation levels of the 23 CpGs that were significantly impacted by any one of the five PFAS, overall (N=151) in the Glowing study, 2010- 2014.

**Figure 3:**
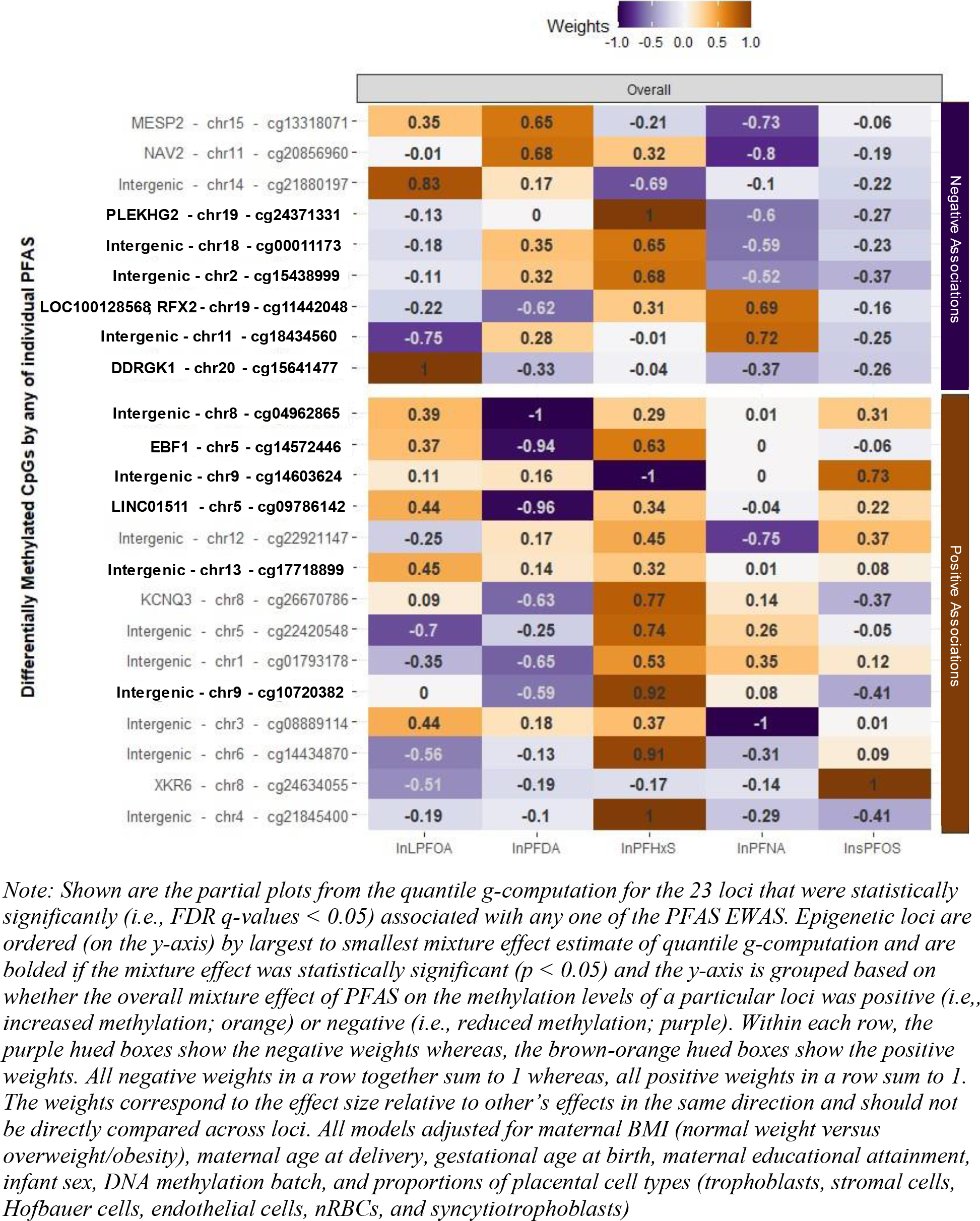
Positive and negative weights showing the partial effects of placental per- and polyfluoroalkyl substances (PFAS in ng/g) on individual placental DNA methylation levels of the 23 CpGs that were significantly impacted by any one of the five PFAS in linear regression models, estimated using quantile g-computation, overall (N=151) in the Glowing study, 2010-2014.

However, multiple interesting findings emerged from the mixture analyses. For instance, PFOS had a negative weight for all loci for which there was a significant negative mixture effect, but other PFAS had stronger negative weights for each of these negative mixture effects. Additionally, PFOS was assigned a positive weight for 4 of the 6 loci with significant positive mixture effects, but only had the strongest weight for cg14603624. Similarly, the PFOA weights aligned with the overall mixture effects for all of the significant mixture effects, except for cg15641477, where it had an opposite direction of effect, and for cg10720382 where it was assigned a weight of zero. The other three PFAS had more variation in their contributions to the significant mixture effects. Interestingly, among the loci with significant mixture effects, only one exhibited the same direction of the weights for all five PFAS, cg17718899. This locus was only significantly associated (FDR < 0.05) with high PFOA levels in the individual EWAS models, but in the mixture model all five PFAS had positive weights, with the strongest positive contributions coming from PFOA, PFHxS, and PFNA, respectively. For most loci, 1, 2, or 3 PFAS were driving the majority of the directional effects, and other PFAS weights were close to zero or in the opposite direction of the mixture effect.

When we tested for interactions between each PFAS included in the mixture, we observed that, a simultaneous increase in all five PFAS by one quartile was linked with reduced methylation at cg20856960 and cg11442048 via potential interactions (**Supplementary Table S2**). Interestingly, both quantile g-computation approaches suggested that methylation levels at cg11442048 (annotated to LOC100128568 and *RFX2* genes on chr19) were reduced upon placental mixture PFAS exposure suggesting that this may be an important target site for several PFAS.

### Sex-Specific Analyses

We explored whether the effect of each individual PFAS on the placental methylome was differential among males and females via stratified analyses (**Supplementary Figure S6** and **Table S1**). We observed that p-values tended to be smaller, effect sizes larger, and larger numbers of statistically significant loci were identified in the sex-stratified models. Specifically, in sex stratified models, we observed that increasing exposure to PFAS tended to be linked with substantially more methylation perturbations among females than males. This was particularly apparent for PFHxS and PFOS levels, which were associated with 644 and 232 loci respectively in females, and with 137 and 125 loci respectively in males (FDR < 0.05).

### Functional Enrichment Analyses

Last, we aimed to understand whether any GO terms or KEGG pathways were enriched among the epigenetic loci that were most differentially methylated with increasing PFAS concentrations. Since the 23 loci that yielded EWAS associations (FDR < 0.05) were only annotated to 10 genes, we expanded our gene list to include any loci that yielded an EWAS association p-value < 1.0e-05, which included 103 loci. No GO terms were significantly enriched after adjusting for bias and multiple testing. One KEGG pathway was significantly enriched (FDR q-value < 0.05), *Valine, leucine and isoleucine biosynthesis* (hsa00290).

## Discussion

We quantified 17 PFAS concentrations in placental tissue from the Glowing cohort of mother-infant dyads in the Central Arkansas region. We show that several placental PFAS concentrations are associated with differential DNAm at several epigenetic loci. In our study, placental PFAS concentrations were moderately correlated with each other. We found that PFHxS exhibited the largest number of significant differentially methylated epigenetic loci. When we compared parameter estimates across EWAS, while still correlated for multiple pairwise comparisons, the parameter estimates for PFHxS had the weakest correlations with the parameter estimates for all other PFAS. Our secondary analyses with mixture modeling revealed that 12 of the 23 tested sites exhibited significant mixture effects and that PFOS and PFOA generally contributed to mixture effects in the same direction, whereas the other three PFAS were more variable in their contributions to mixture effects. Additionally in the mixture analyses, PFHxS was assigned strong weights in opposite directions of other PFAS for many of our identified loci. These findings could suggest that PFOS and PFOA may have more synergistic or cumulative impact on the placental methylome and that PFHxS has a more unique effect on the placental epigenome compared to the other 4 PFAS that we tested. Notably, PFHxS has been a common replacement for PFOS and PFOA, under the assumption that shorter chain PFAS may be less toxic than legacy PFAS [23]. PFHxS was also very highly detected in the ground water at an air force base north of Little Rock, AR [24]. Thus, the potential impacts of prenatal PFAS exposure, and particularly PFHxS, are important for this and similarly exposed populations.

The vast majority of epidemiologic studies have examined concentrations of PFAS in maternal samples, while only a handful have quantified placental PFAS concentrations, which may be more reflective of direct *in utero* exposure. We compared our placental PFAS levels to one other published study from a US cohort, where PFAS were quantified in placenta sampled between 2010 – 2011 in Durham, North Carolina. Complimentary to our findings, Hall et al. (2022) observed similar average placental concentrations of 4 PFAS, a similar correlation structure among these PFAS, and that PFOS had highest concentrations [25]. While Hall et al. did not examine placental epigenetic responses, they did find that increasing concentrations of placental PFAS were associated with smaller birth size for gestational age and that there may be some sex-specific effects on fetal growth [25].

Interestingly, several of the genes and epigenetic loci that we identified as differentially methylated in association with increasing placental PFAS concentrations have been associated with other endocrine disrupting chemicals and with cardiometabolic outcomes. For instance, multiple CpGs within *XKR6*, among other CpGs and genes, were found to have lower DNA methylation levels among placenta of newborns with selective fetal growth restriction [26]. Higher levels of maternal urinary bisphenol A (BPA) concentrations were associated with lower DNA methylation of one CpG in *XKR6* [27].

Additionally, maternal glucose concentrations during early pregnancy were associated with differences in DNA methylation at one CpG in *XKR6* [28]. Placental DNA methylation levels of CpGs in *NAV2* have previously been associated with maternal systolic blood pressure [29, 30]. The expression of *KCNQ3* was shown to be upregulated in placentas from mother with preeclampsia [31]. Notably, *Early B Cell Transcription Factor 1* (*EBF1),* annotated to cg14572446, has been associated with multiple traits (GWAS catalog [32]), including anthropometrics, body composition, and lipid profiles, and CpGs annotated to *EBF1* were also identified in the only other placental PFAS-EWAS to date [9]. Interestingly, in our study, DNAm of cg14572446 within *EBF1* was positively related to simultaneous increase in both PFHxS and PFOA. *In-vitro* experiments showed that Bisphenol S (BPS) was associated with decreased expression of *MESP2* in preadipocytes [33]. While our functional enrichment analyses only yielded one significantly enriched KEGG pathway – *valine, leucine and isoleucine biosynthesis*, it is of note that several untargeted metabolomics studies of PFAS have identified this same pathway being perturbed with PFAS exposure [34]. Prenatal exposure to PFAS has been associated with non-alcoholic fatty liver disease in children, and perturbations to valine, leucine, isoleucine metabolism and biosynthesis, along with other amino acids and lipids, were identified as potential intermediaries [35, 36]. In another study, maternal plasma concentrations of PFAS were associated with perturbations to multiple amino acid metabolism and biosynthesis pathways, with leucine, isoleucine, valine metabolism exhibiting the greatest number of associations [37]. Circulating levels of valine, leucine, and isoleucine are also important predictors or biomarkers of several cardiometabolic diseases [38]. Overall, many of the genes annotated to our PFAS-associated epigenetic loci serve important roles in cardiometabolic health, and some of these have been shown to be affected by other endocrine disrupting chemicals.

In addition to cardiometabolic health, some of these genes are important in neurodevelopment. In a rat model, prenatal BPA exposure was associated with altered expression of *KCNQ3* in the amygdala of exposed females [39], and *KCNQ3* may have important functions in some neurodevelopmental outcomes [40]. *NAV2* guides axonal outgrowth and plays critical roles in early cerebellar development [41], while *PLEKHG2* is also involved in axonal growth, and impaired function is likely involved in neuronal disorders [42]. Importantly, our study did not examine whether DNAm at our PFAS-associated epigenetic loci was associated with the expression of the genes or metabolites that we describe above. Additionally, many genes have pleiotropic effects with roles in numerous biological processes and phenotypes. The actual genomic and phenotypic consequences of our observed differential DNAm are not clear, but these prior studies provide important context about the biological process and potential phenotypic outcomes that may be involved in such responses to prenatal PFAS exposures.

Our study contributes to the emerging evidence that PFAS impact the epigenetic regulation of the placenta and these findings may have implications for cardiometabolic health or other children’s health outcomes. Importantly, our study utilized placental PFAS concentrations to study the impacts on the placental methylome, allowing for a more proximal exposure biomarker that may also be highly impactful on the *in-utero* environment. However, there are several limitations of this work that should be considered when interpreting our findings. While we utilized two data reduction steps to improve power and applied an FDR correction to identify significant differential DNAm, our sample size was modest for an epigenome wide approach (n=151). This moderate sample size also resulted in reduced statistical power, particularly in the sex-stratified analyses. However, our sex-stratified analyses are hypothesis-generating, and suggest that future studies of placental PFAS and DNAm should explicitly plan for and perform analyses to identify sex-specific effects. Additionally, the Glowing Cohort represents a healthy population in central Arkansas. While this reduces potential generalizability to other populations, our study also represents an important contribution to the characterization of risk factors for adverse child growth and cardiometabolic health in a population that has potentially high exposure to some PFAS and high rates of low birthweight. Despite some limitations, our study provides a substantial contribution towards understanding the placental epigenetic responses to PFAS.

## Conclusions

We performed an EWAS of placental PFAS levels associated with placental DNAm in a healthy cohort of mother-infant dyads from Arkansas. We found that numerous epigenetic loci in the placenta appear to be perturbed in association with increasing PFAS concentrations, and that PFHxS had the most pronounced effect in this study. Mixture analyses revealed that PFOA and PFOS tended to have consistent directions and possibly cumulative effects on several epigenetic loci, while PFHxS tended to perturb the placental methylome more independently. Additionally, we provide suggestive evidence that sex-specific associations are an important consideration for future studies. The genes that are annotated to our identified loci predominantly play important roles in growth and cardiometabolic health, but we also identified several genes that are important in neurodevelopment. These findings can provide insights into the mechanisms through which prenatal PFAS exposures impact birth outcomes and children’s health.

## CRediT authorship contribution statement

**Todd M Everson**: Conceptualization, Data curation, Formal analysis, Funding acquisition, Methodology, Writing – original draft, Writing – review & editing. **Neha Sehgal**: Formal analysis, Methodology, Visualization, Writing – original draft, Writing – review & editing. **Dana Boyd Barr**: Data curation, Methodology, Writing – review & editing. **Parinya Panuwet**: Formal analysis. **Volha Yakimavets**: Formal analysis. **Cynthia Perez**: Formal Analysis. **Stephanie M. Eick**: Methodology, Writing – review & editing. **Karitk Shankar**: Formal Analysis, Methodology, Data Curation, Writing – review & editing. **Kevin J Pearson**: Funding Acquisition, Project administration, Writing – review & editing. **Aline Andres:** Data Curation, Program Administration, Funding acquisition, Writing – review & editing.

## Declaration of competing interest

The authors declare that they have no known competing financial interests or personal relationships that could have appeared to influence the work reported in this paper.

## Data Availability

All data produced in the present study will be made available upon publication.

## Acknowledgments

We acknowledge all the families who chose to participate in this study as well as the Arkansas Children’s Nutrition Center team for the data and samples acquisition for the Glowing Study.

## Funding statement

This work was supported by funding from the National Institute of Environmental Health Sciences, NIH/NIEHS [R01 ES032176; MPIs: Andres, Everson, & Pearson], the HERCULES Center [P30 ES019776; Everson, Barr, Eick] and the USDA-ARS [6026-51000-012-06S; Glowing cohort].

Disclaimer: The content is solely the responsibility of the authors and does not necessarily represent the views NIH.

## Data availability

Raw and processed epigenetic data will be made available on NCBI GEO upon publication.

## Notes

### Competing Interest Statement

The authors have declared no competing interest.

### Author Declarations

All study participants provided informed, written consent and the study was approved by the University of Arkansas for Medical Sciences Institutional Review Board.

