## Supplemental Tables and Figures. for "Placental PFAS concentrations are associated with perturbations of placental DNA methylation at loci with important roles on cardiometabolic health"

**Supplementary File for “Placental PFAS concentrations associated with perturbations of placental DNA methylation”**

**Supplemental Figure S1: Direct Acyclic Graph (DAG) for the association between placental DNA methylation and placental per- and polyfluoroalkyl substances (PFAS)** **exposure in the Glowing study, 2010-2014.**

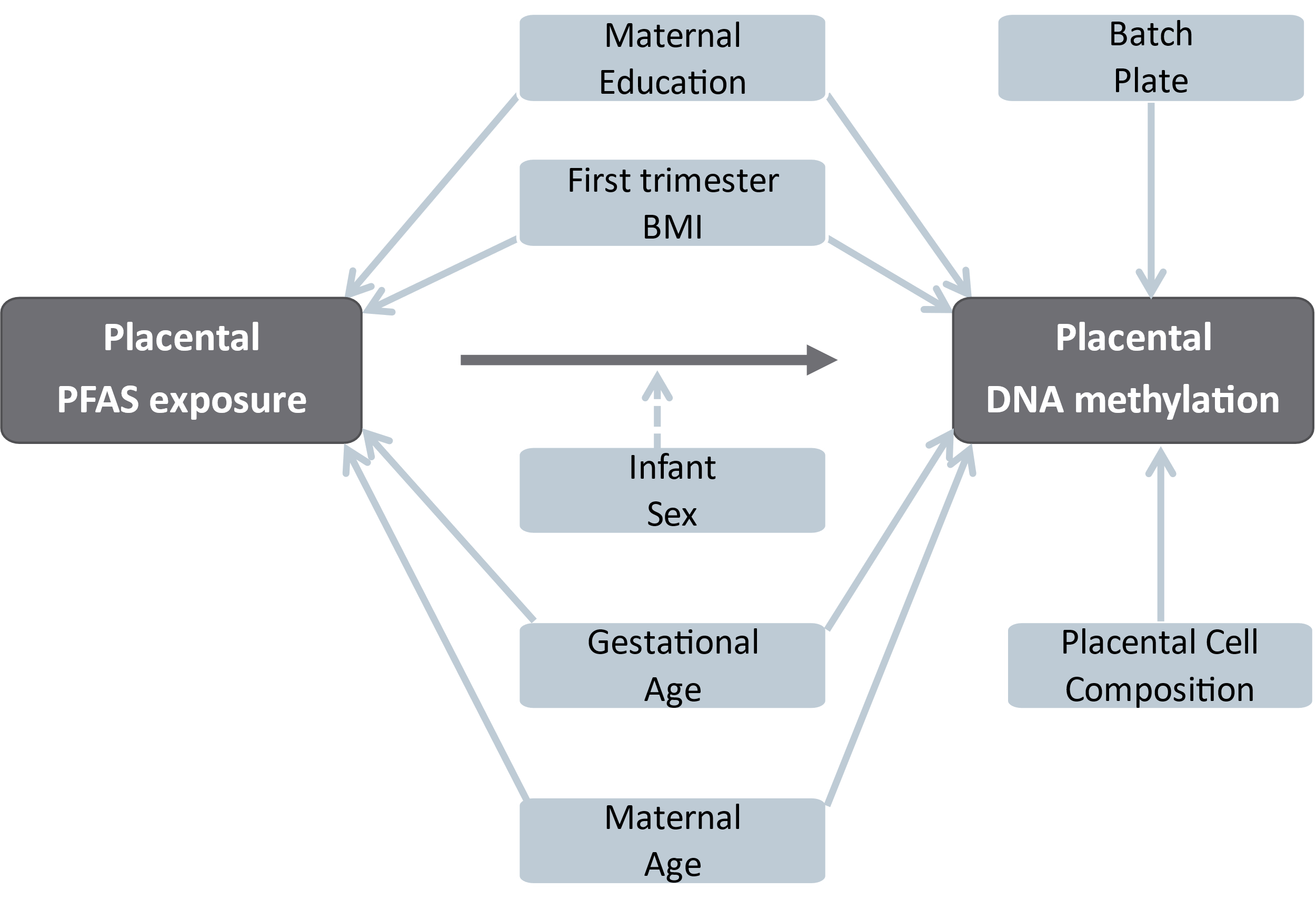

**Supplemental Figure S2: Violin plots for the distribution of natural log transformed PFAS concentrations from placental tissues in the Glowing study, 2010-2014.**

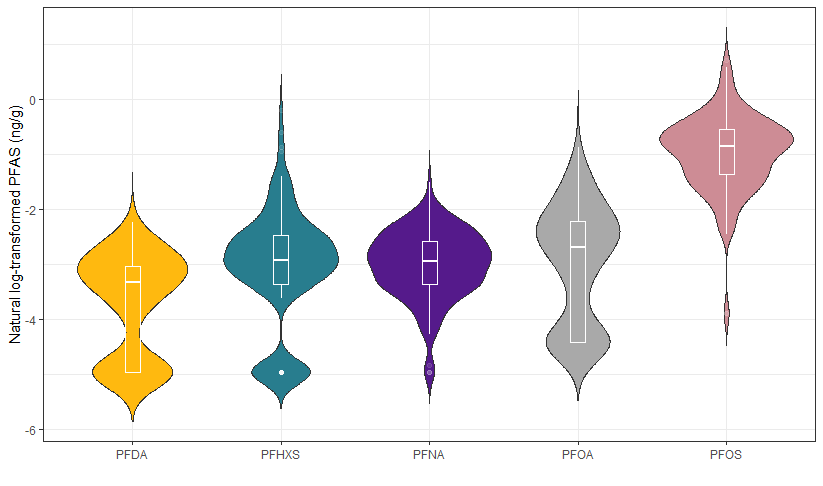

**Supplemental Figure S3: Spearman correlation of natural log-transformed placental per- and polyfluoroalkyl substances levels (ng/g) in the Glowing study, 2010-2014.**

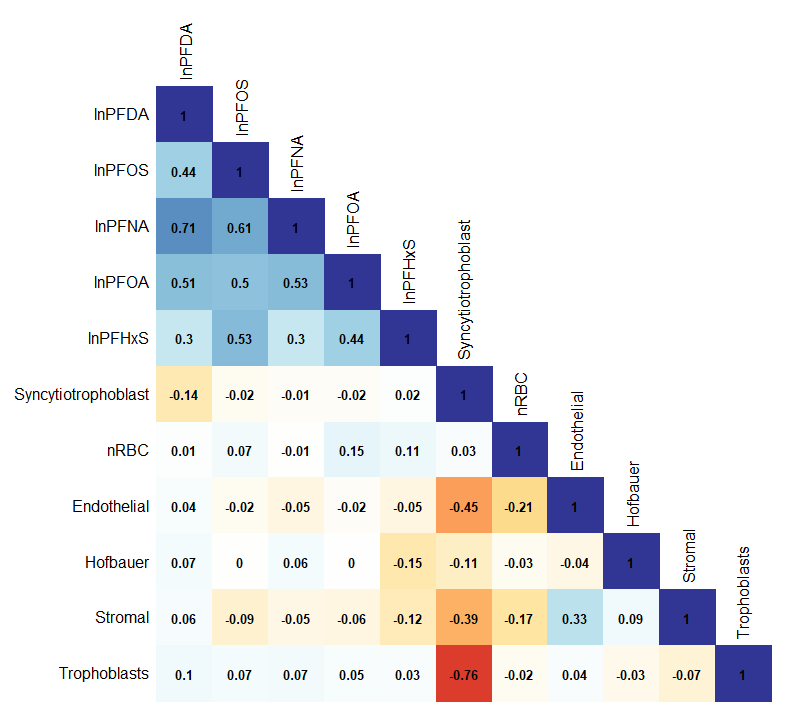

**Supplemental Figure S4. QQ Plots for EWAS of PFOS (λ = 0.94), PFOA (λ = 0.99), PFHxS (λ = 1.15), PFDA (λ = 1.09), and PFNA (λ = 0.86).**

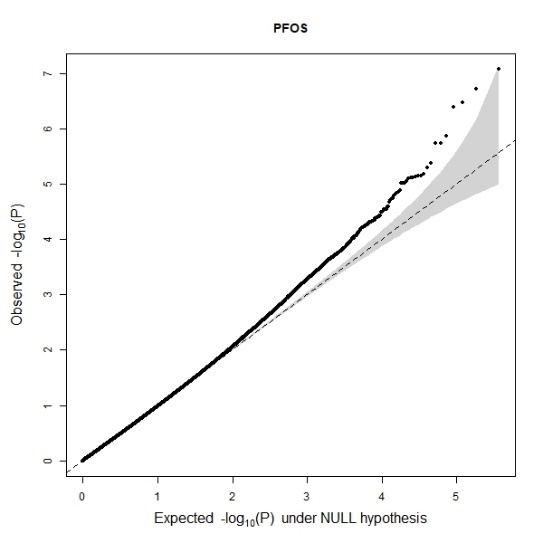

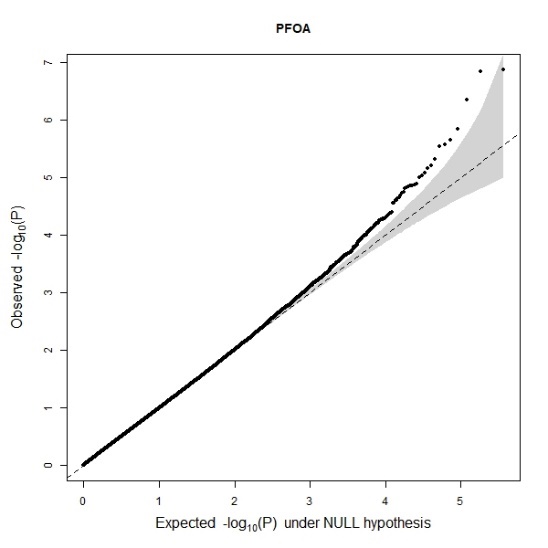

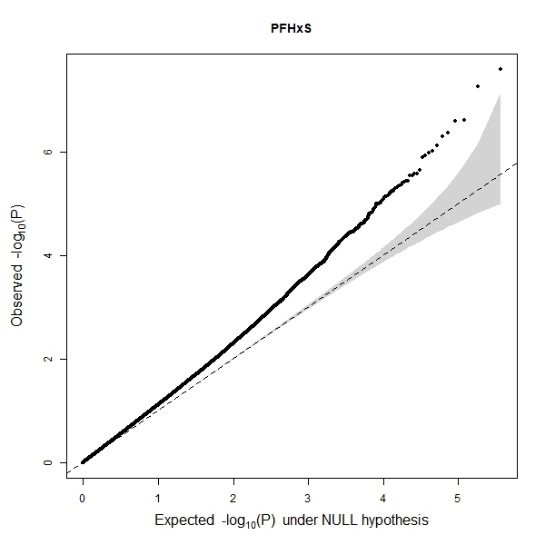

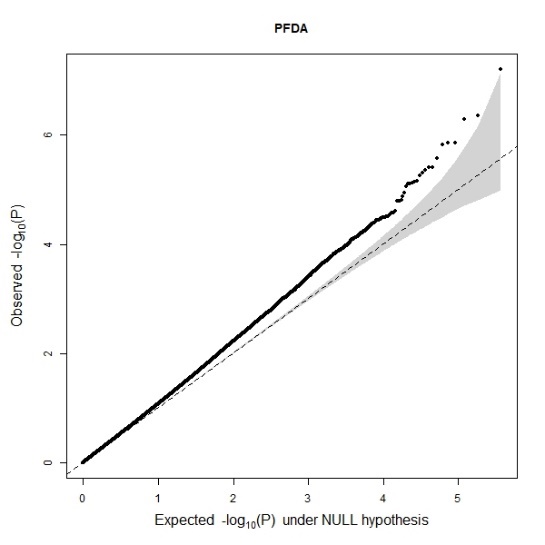

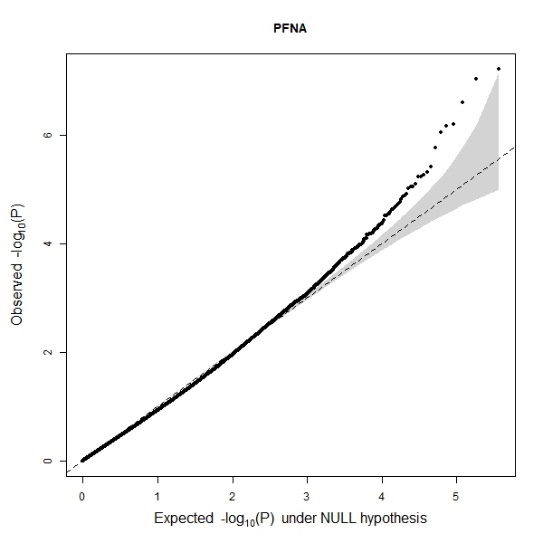

**Supplemental Figure S5. Bivariate scatter plots (below the diagonal), distributions (on the diagonal), and Spearman correlation rho values (above the diagonal) between EWAS parameter estimates (x- and y-axes) for the 23 CpGs that were associated with one of PFOS, PFOA, PFHxS, PFDA, or PFNA.**

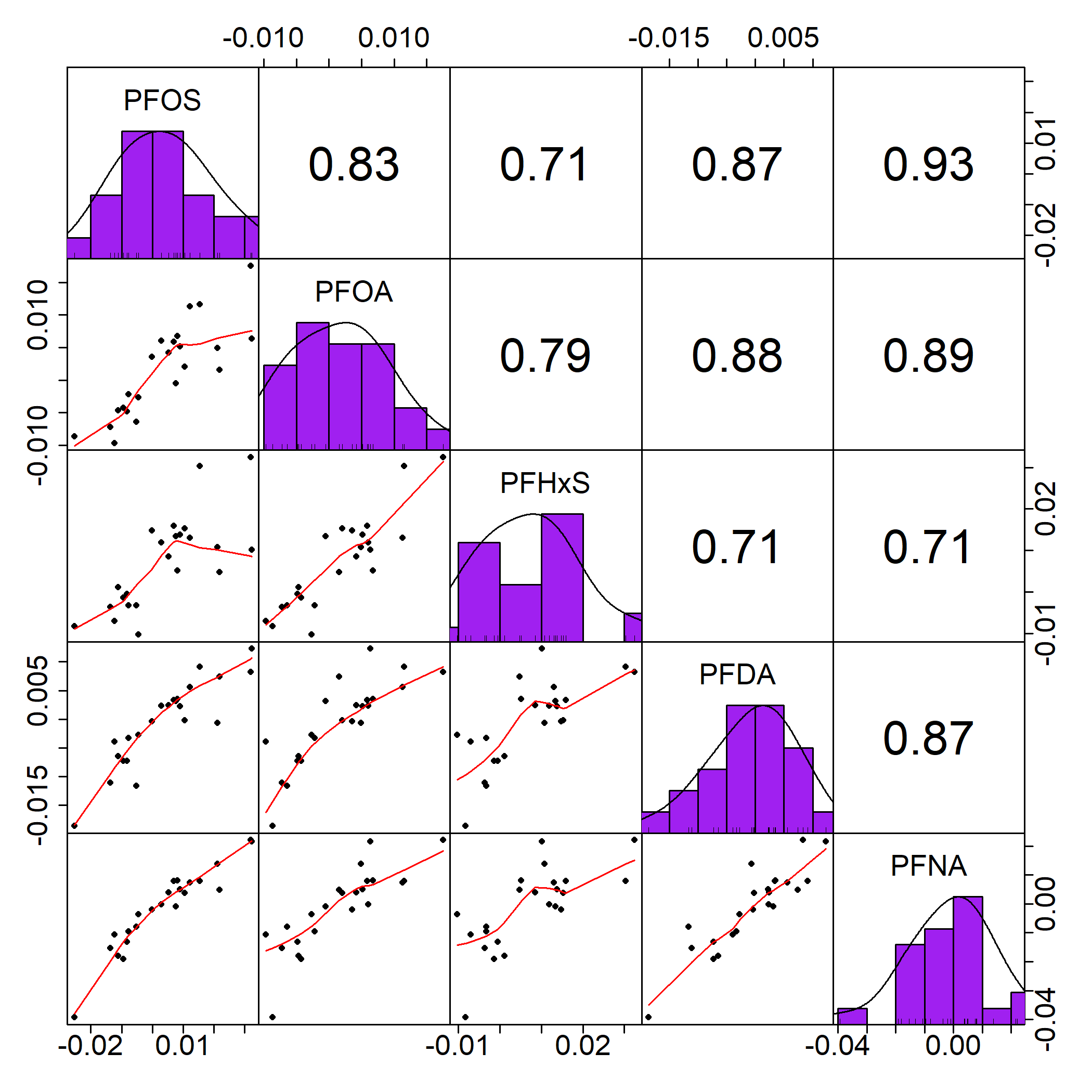

**Supplemental Figure S6: Volcano plots for the epigenome wide-association between placental per- and polyfluoroalkyl substances levels (ng/g) and placental DNA methylation of the 365,376 highly methylated CpGs, overall (N=151) and among only females (N=63) and males (N=88) in the Glowing study, 2010-2014.**

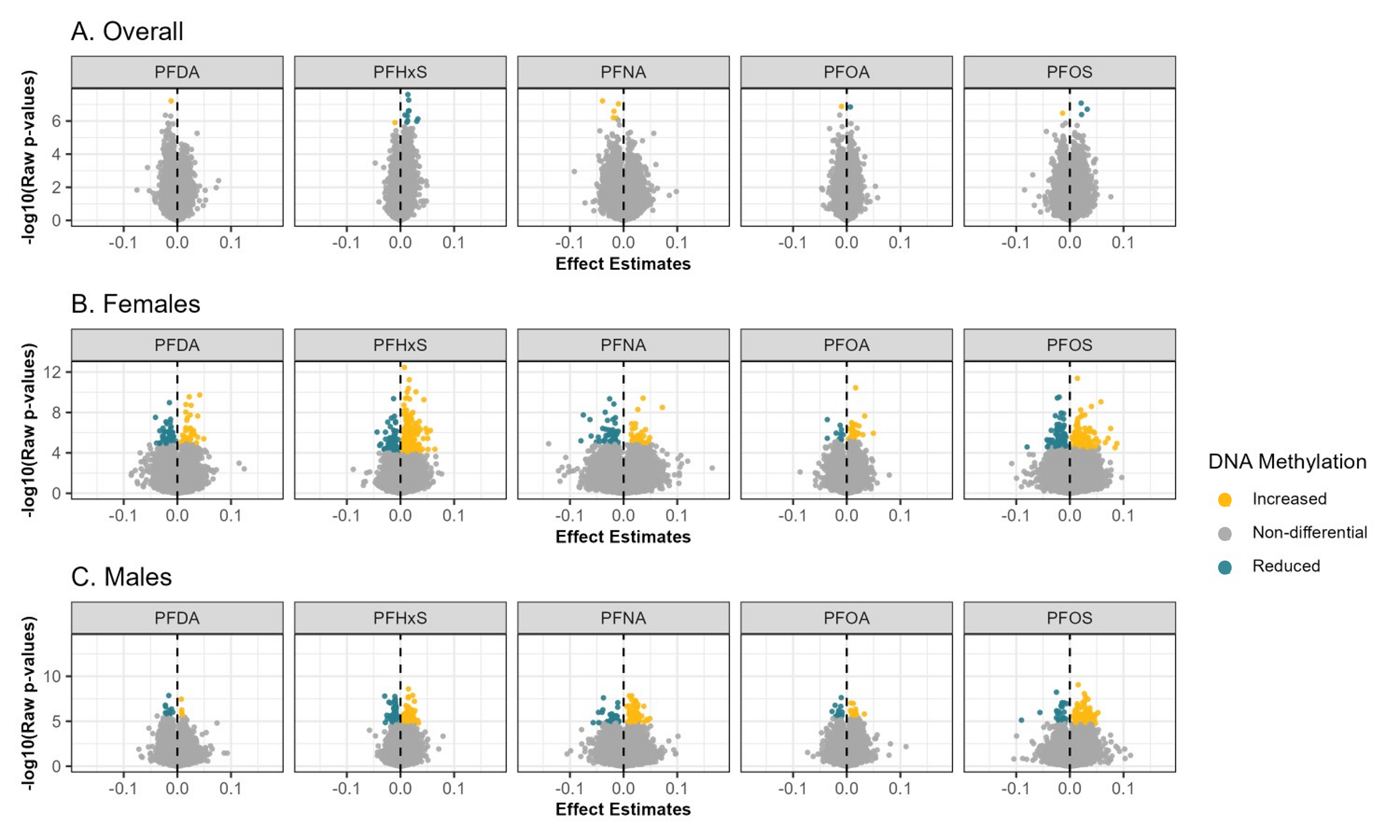

*Note: All models adjusted for matneral BMI (normal weight versus overweight/obesity), maternal age at delivery, gestational age at birth, maternal educational attainment, infant sex, DNA methylation batch, and proportions of placental cell types (trophoblasts, stromal cells, Hofbauer cells, endothelial cells, nRBCs, and syncytiotrophoblasts). Increased or reduced methylation is based on whether the association was above or below 0, respectively and the FDR q-values < 0.05.*

**Supplemental Table S1. Summary results of epigenome wide association studies (EWAS) for the relationship between placental per- and polyfluoroalkyl substances levels (ng/g) and placental DNA methylation, overall (N=151) and among females (N=63) and males (N=88) only in the Glowing study, 2010-2014.**

| Exposure | Infant Sex | N | CpGs with reduced methylation with higher PFAS | CpGs with increased methylation with higher PFAS | Inflation factor |
| --- | --- | --- | --- | --- | --- |
| PFOA | Overall | 151 | 1 | 1 | 0.99 |
|  | Females | 63 | 10 | 35 | 0.90 |
|  | Males | 88 | 14 | 12 | 1.07 |
| PFDA | Overall | 151 | 1 | 0 | 1.09 |
|  | Females | 63 | 49 | 38 | 0.97 |
|  | Males | 88 | 11 | 4 | 0.99 |
| PFHxS | Overall | 151 | 1 | 10 | 1.15 |
|  | Females | 63 | 75 | 569 | 1.14 |
|  | Males | 88 | 66 | 71 | 1.15 |
| PFNA | Overall | 151 | 5 | 0 | 0.86 |
|  | Females | 63 | 50 | 30 | 0.90 |
|  | Males | 88 | 35 | 76 | 0.89 |
| PFOS | Overall | 151 | 1 | 3 | 0.94 |
|  | Females | 63 | 114 | 118 | 0.93 |
|  | Males | 88 | 28 | 97 | 0.97 |

*Note: In the five individual, untargeted EWAS analyses, CpGs had increased or reduced methylation if their associations were above or below 0 respectively and statistically significant (FDR q-values < 0.05). All models adjusted for maternal BMI (normal weight versus overweight/obesity), maternal age at delivery, gestational age at birth, maternal educational attainment, infant sex, DNA methylation batch, and proportions of placental cell types (trophoblasts, stromal cells, Hofbauer cells, endothelial cells, nRBCs, and syncytiotrophoblasts).*

**Supplemental Table S2. Summary results of mixture effects of PFAS levels (ng/g) and placental DNA methylation, overall (N=151) and among females (N=63) and males (N=88) only in the Glowing study, 2010-2014.**

| *Targeted, mixture analysis using quantile g-computation* | Model | Infant Sex | N | CpGs with reduced methylation with increasing PFAS | CpGs with higher methylation with increasing PFAS |
| --- | --- | --- | --- | --- | --- |
| No interaction | 23 individual CpGs ~ mixture of lnPFAS + covariates | Overall | 151 | 6 | 6 |
|  |  | Females | 63 | **1** | 0 |
|  |  | Males | 88 | **5** | 3 |
| Allowing for interaction | 23 individual CpGs ~ mixture of lnPFAS + I(PFOA*PFNA*PFHxS*  PFOS*PFDA) + covariates | Overall | 151 | **2** | 0 |
|  |  | Females | 63 | 0 | **1** |
|  |  | Males | 88 | **2** | 0 |

*Note: CpGs had increased or reduced methylation if their associations were above or below 0 respectively and statistically significant (p-values<0.05). All models adjusted for maternal BMI (normal weight versus overweight/obesity), maternal age at delivery, gestational age at birth, maternal educational attainment, infant sex, DNA methylation batch, and proportions of placental cell types (trophoblasts, stromal cells, Hofbauer cells, endothelial cells, nRBCs, and syncytiotrophoblasts).*
